# Development and Validation of a Nutrient Profiling Model for Shopping Baskets: The Grocery Basket Score (GBS) Methodology

**DOI:** 10.1101/2025.01.03.25319947

**Authors:** Paul Windisch, Sandro Marcon, Javier Orts, David Faeh, Richard Hurrell, André Naef

**Affiliations:** dacadoo AG, Zurich, Switzerland; Epidemiology, Biostatistics and Prevention Institute, Division of Chronic Disease Epidemiology, University of Zurich, Zurich, Switzerland; Health Department, Bern University of Applied Sciences, Bern, Switzerland; Professor emeritus, Institute of Food, Nutrition and Health, ETH Zurich, Switzerland

**Author notes:** **Correspondence:** Paul Windisch, MD dacadoo AG Othmarstrasse 8 8008 Zurich.

## Abstract

**Background:** Rating a person’s diet as a whole, as opposed to rating individual food items, could help better inform consumers about the health value of their diet. Our goal was to develop an automated health rating of grocery shopping baskets, based on the nutrient composition of all the food items contained within the basket, and without knowing how many people consume the food items over what period of time. It is envisaged that the rating, or score, can be deployed by retailers on top of existing loyalty or digital shopping basket programs.

**Methods:** We developed a novel model for calculating Grocery Basket Scores (GBS) that uses nutrient energy densities rather than absolute quantities. It was based on self-reported daily food intake from the National Health and Nutrition Examination Survey (NHANES) as well as mortality follow-up data. We conducted a validation against the Alternate Healthy Eating Index (AHEI) as well as the Nutri-Score.

**Results:** The nutrient energy density (GBS) model penalized consuming calories from sugar and saturated fats, high salt intake, as well as consuming calories from beverages. Furthermore, a penalty was applied to foods that have a low ratio of protein, vitamin C, or iron relative to the total number of calories. Fiber consumption was rewarded. The model showed a high degree of correlation with the AHEI (absolute Pearson correlation coefficient: 0.62 for the AHEI without the protective effect of moderate alcohol consumption and 0.60 with it) and the Nutri-Score (absolute Pearson correlation coefficient: 0.60).

**Conclusions:** The proposed nutrient energy density model is aligned with the recommendations formulated in nutritional guidelines, as indicated by a high degree of correlation with the AHEI and Nutri-Score. Long term use of a GBS from a single or multiple grocery stores could help consumers adhere to dietary guidelines.

## Introduction

Although the impact of diet on non-communicable diseases and mortality has been investigated in many studies (1), what constitutes an optimal diet remains subject to discussion. As a consequence, nutrition profiling and rating systems have been developed to inform consumers and nudge them towards healthier shopping choices (2).

One example is the Nutri-Score, which has been endorsed by several countries and is found on a variety of products (3). Profiling systems such as the Nutri-Score focus on rating individual foods within a given group, which allows for printing scores on the products and enables the consumer to make direct comparisons between different items from the same category (4). While this is useful, focusing on individual foods also comes with tradeoffs. For example, one could compose an entire shopping basket consisting exclusively of items that received an A grading by the Nutri-Score due to their low-calorie density and high fiber content while containing little protein, even though the Nutri-Score rewards protein. As protein deficiency has now become an issue in the elderly in industrialized nations (5) it illustrates that assessing nutrition as a whole is more complex than summing up or even averaging individual food ratings. This is also relevant to consumers, as they must decide what share of poorly rated products in their basket is still acceptable.

Nutritional guidelines such as the Alternate Healthy Eating Index (AHEI) tend to provide recommendations in quantities, such as servings, per day (6,7). The AHEI was designed to predict chronic disease risk and has also been validated for predicting mortality (8). While this approach can help consumers understand how they should compose their diet, it can still be challenging to translate this information into actionable shopping decisions as a consumer must still assess how many servings are contained in a single product and keep track of the number of products they need for a given timeframe.

Our goal was to develop a nutrient energy density model that avoids serving sizes and can be used directly in supermarkets to translate recommendations from nutritional guidelines into actionable decisions and, at the same time, will complement product-level rating systems such as Nutri-Score. To this end, we set out to develop a model to rate shopping baskets for their health properties, assuming that these baskets account for a substantial amount of a consumer’s total nutrition. Although a single basket of food products may have little relevance, long term purchases from one retailer, or from a group of retailers who have accepted to share information, would be meaningful.

Additionally, we did not want to develop a model that required a manual ingredient annotation but instead wanted a model that could be automatically deployed by retailers on top of existing loyalty or digital shopping basket program. Finally, we sought a data-based model that avoided strong reliance on opinionated expert judgment and based our nutrient energy density model on the link between nutrition and eventual mortality. We conducted a validation by comparing the outputs of the final GBS model with the AHEI and the Nutri-Score to ensure seamless integration into existing guidance.

The hypothesis was that while there would be differences with the Nutri-Score due to the factors mentioned above, GBS and Nutri-Score would be in agreement for the majority of items. For the AHEI, we hypothesized a better agreement with GBS than with Nutri-Score, due to the alignment of rating the consumer’s food purchases as a whole (i.e. dietary patterns) as opposed to rating individual items.

## Methods

### Data

The data used in this study is a subset of the continuous National Health and Nutrition Examination Survey (NHANES) (9) from 2001 to 2018, which has been linked with the latest mortality data for the participants from 2019 (10). The dietary data thus does not constitute data derived from retail purchases, but is self-reported complete daily food intake data from each participant. As part of the NHANES nutritional assessment, participants of the respective cohort completed a 24-hour dietary recall interview in person with a trained dietary interviewer and participated in a second phone-based interview 3 to 10 days later. The food intake obtained from these recalls was then linked through codes to the Food and Nutrient Database for Dietary Studies (FNDDS) (11),and was used to calculate the respective daily nutrient intakes for each participant in each recall.

Individuals younger than 17, older than 90, those with missing nutritional information, and those who reported no calorie intake, were excluded from the analysis. This resulted in a dataset containing the daily nutrient intake of 35’462 individuals. Data from 25% of the individuals was randomly assigned to the test set, with data from the remaining individuals being used as the training set to model eventual mortality from dietary signals.

### Considerations for Potential Dietary Signals

We generated two types of dietary signals, namely “nutrient energy density signals” and “low nutrient-energy-ratio signals”. As one cannot know how long a shopping basket will last in a household, or how many people consume the food items in the basket, we calculated dietary signals based on energy density, that is energy or weight from a given nutrient relative to the total basket energy.

Nutrient energy density signals that were evaluated over the course of the project were calories from carbohydrates divided by total calories, calories from sugar divided by total calories, calories from proteins divided by total calories, calories from fat divided by total calories, calories from saturated fats divided by total calories, calories from unsaturated fats divided by total calories, calories from alcohol divided by total calories, amount of fiber divided by total calories, amount of sodium divided by total calories, and calories from beverages divided by the weight of the beverages.

A further set of signals were calculated for proteins and certain vitamins and minerals as indicators of low dietary intake. These “low nutrient-energy-ratio signals” indicate that a person is consuming a low amount of a given nutrient relative to their total calorie consumption. In order to calculate the nutrient-energy-ratio signal, the daily recommended intake of the nutrient was divided by the daily recommended calories, resulting in a recommended density of the nutrient per calorie. The low nutrient-energy-ratio signal was included in the model if the nutrient per calorie density was below the recommended density. The signal becomes stronger the lower the density gets. The nutrients which were evaluated as low nutrient-energy-ratio signals and the respective recommendations that were used for the calculation are provided in Supplementary Table 1. Nutrient-energy-ratio signals were generated for vitamin C, folate, potassium, calcium protein, and iron.

All dietary signals were then evaluated as potential inputs to Cox proportional hazard models for predicting mortality. As a requirement for all dietary signals, there should be a plausible biological mechanism linking each signal to mortality. Such a link should be supported by a consistent correlation of the respective signal with mortality in NHANES (9).

Another consideration was that the signals should be computable for any retailer without additional manual labeling of foods being required. Therefore, we focused on nutrients that are either part of the labeling requirements in many countries or that could be reliably imputed, e.g., from the name of the food and its categories. Lastly, we wanted the model to be based on different, heterogeneous signals to reduce the likelihood of single items achieving very high scores by performing well across all signals, as a shopping basket consisting only of one item is unlikely to be conducive to healthy nutrition (low dietary diversity). In other words, we sought a mix of signals that provides a reasonable fingerprint of grocery shopping behavior.

### Modeling

A schema of the modeling process is presented in Figure 1. For the modeling and validation, we considered the items from the daily dietary recall as a person’s shopping basket. Due to high degrees of correlation between different potential signals, different combinations of signals were evaluated while omitting others. The goal was to minimize redundancy, identify combinations of signals that fulfill all requirements mentioned above, avoid multicollinearity as a source of model misspecification, and ultimately to use the signals as inputs to a Cox proportional hazard model. Avoiding multicollinearity was particularly important as many potential signals were highly correlated (e.g. calories from sugar divided by total calories, calories from added sugar divided by total calories, and calories from carbohydrates divided by total calories).

**Figure 1:**
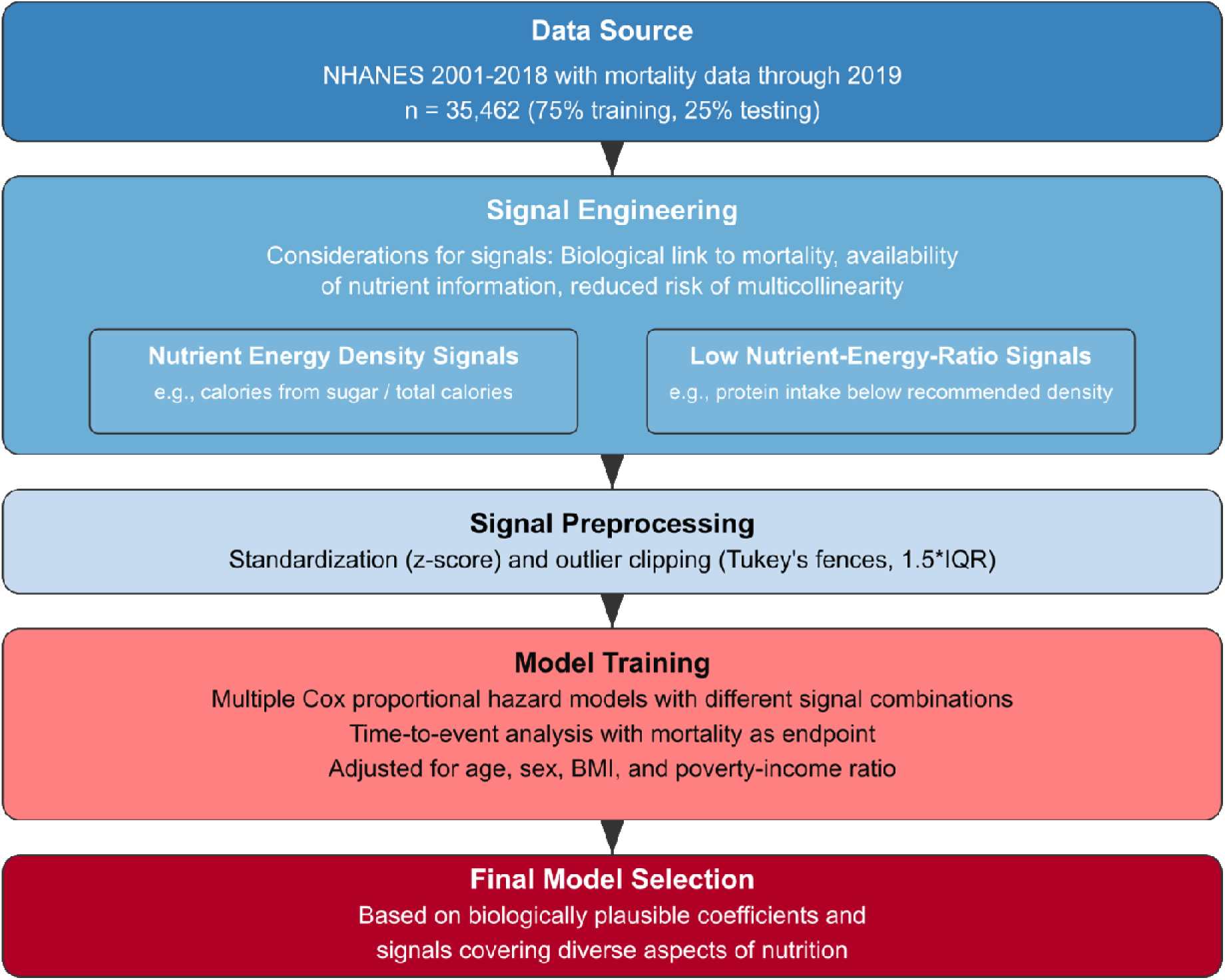
Schema of the Grocery Basket Score (GBS) Modeling Process. Conceptual framework for the development of the Grocery Basket Score using NHANES 2001-2018 data linked with mortality data (n=35,462). Following data preprocessing and standardization, two categories of dietary signals were generated: nutrient energy density signals and low nutrient-energy-ratio signals. Signal construction was informed by established biological mechanisms, nutrient information availability, and multicollinearity considerations. Multiple Cox proportional hazard models, adjusted for demographic and anthropometric covariates, were evaluated. The final GBS model was selected based on coefficient plausibility and comprehensive representation of diverse nutritional aspects.

If highly correlated signals are used as independent variables in a regression, the model cannot reliably separate them, resulting in wild coefficient swings with small changes in the data. This increases the variance of the estimates and leads to wider confidence intervals. Moreover, as multicollinear variables tend to "cancel out", they can lead to coefficients that are not bio-plausible when viewed in isolation, thus making the interpretation of the model difficult, as expected relationships between dietary patterns and mortality risk disappear. In addition, we evaluated a joint model that used the same signals for both foods and beverages as well as separate models with different signals for foods and beverages respectively.

Standardization of the signals was performed for interpretability reasons, by subtracting the mean and dividing by the standard deviation, such that the resulting model coefficients are on the same scale. Signals were also clipped to the limits of 1.5 times the interquartile range (IQR) rule for outlier detection (Tukey’s fences).

Since the model’s goal was to predict a time-to-event endpoint and the NHANES data is censored, Cox proportional hazard models were used to estimate the effect of different combinations of the dietary signals (defined above) on mortality. The models were adjusted for age, sex, body-mass index (BMI) and the poverty-income ratio (i.e. household income divided by the poverty threshold) by including them as model coefficients and setting them to zero at inference time. The Cox model which included a comprehensive set of dietary signals and showed good performance at predicting mortality, while including a comprehensive set of signals, was chosen for further validation. This nutrient energy density model was then used as the basis for the Grocery Basket Score (GBS).

The output that is obtained by multiplying the regression coefficient with the strength of the respective dietary signal is the logarithm of the hazard ratio. Afterwards, the hazards were transformed in order to achieve a score resembling a normal distribution ranging from zero (worst) to one thousand (best) which is how the score was supposed to be presented to users.

To this end, the obtained values were clipped beyond -0.8 and 1.5, normalized between 0 and 1, and subtracted from 1. Values were then raised to a power of 1.5. and multiplied by 1’000.The separate levels of adjustment for the core and beverages model are available in Supplementary Tables 2 and 3, respectively. Unless specified otherwise, data analysis was performed using Python (v. 3.8.10) as well as the numpy (v. 1.23.5), pandas (v. 1.3.5) and lifelines (v. 0.27.7) packages.

### Validation

For the validation against the AHEI, we computed the AHEI scores for 4’382 subjects with dietary recalls from NHANES 2017-18, computed the GBS using the same recalls and calculated the correlation coefficient between the GBS and AHEI, both with and without the protective effect of moderate alcohol consumption. The AHEI was computed in R (v. 4.3.2) using the dietary index package (v. 1.0.3). In addition, we also computed the Nutri-Score 2017 using the pyNutriScore package (v.0.0.2) and calculated a total Nutri-Score for each participant’s nutritional information as a calorie-weighted average of the Nutri-Scores of the individual food items they consumed. We then calculated the correlation between the calorie-weighted Nutri-Scores and the AHEI.

For the validation against the Nutri-Score, we retrieved food items from the Open Food Facts database (12). In the preprocessing, we ignored items with a missing product code or name. Furthermore, we filtered items with missing energy information, missing Nutri-Score 2023, or where the macronutrients summed to more than 101 grams per 100 grams of food, items with saturated fat contents larger than total fat, items with more sugar than carbohydrates, items where one of the nutrients is larger than the 99^th^ percentile and items missing information regarding any of protein, fat, carbohydrate, or salt content. The remaining 913’298 items were scored using the GBS model and compared to the provided Nutri-Scores by computing the Pearson correlation coefficient.

Due to occasionally missing labeling information in the Open Food Facts database, we also developed an imputation model to complete the required label information. Specifically, our model is a deep neural network that implements a sequence model operating on the product categories and name using custom embeddings to impute fiber, iron, and vitamin C. A database with the imputed label information is available at https://github.com/windisch-paul/OFF-GBS.

In addition to computing the correlation coefficient for the whole dataset, clustering was performed to allow for a cluster-by-cluster correlation and comparison. Items were clustered into different food clusters based on their assigned tags in the Open Food Facts database. Due to the high number of tags in the database and each item potentially being assigned multiple tags, it was deemed infeasible to manually define disjointed food clusters. A clustering approach based on context frequency of tags was used instead.

As a first step, we filtered out tags that appeared fewer than 10’000 times in the whole corpus. This left 78 tags. As a second step, the term frequency-inverse document frequency matrix (tf-idf) was calculated. The idea was to vectorize tags based on their frequency and relevance. The tf-idf matrix was then reduced using singular-value decomposition (SVD) by keeping the 19 most relevant components. Finally, the K-Means clustering algorithm was applied to find 14 food clusters, with the 15^th^ cluster manually assigned to beverages. The rationale of performing SVD is that K-Means would perform better on the reduced eigenvector space rather than on the original tf-idf matrix. The number of singular values and clusters was selected empirically using the “elbow-rule”.

In addition to computing correlation coefficients, we calculated the absolute number and the percentage of items in a food cluster for which the difference between the z-scores of the GBS and the Nutri-Score was greater than 1, as well as the number and percentages of items with a difference greater than 2. This was done to highlight the items that caused the biggest disagreement.

## Results

### Modeling

During follow-up, 14.6% (n = 5’167) of participants died (training set: 3’839 deaths out of 26’598 participants = 14.4%; test set: 1’328 deaths out of 8’874 participants = 15.0%). After several iterations, an architecture obtained by scoring the combination of foods and beverages using a core model, and only the beverages using a dedicated beverages model then, combining the results as a calorie-weighted average of the respective scores, demonstrated areas under the receiver operating characteristic curve (ROC-AUCs) ranging from 0.854 for estimating mortality over a 1-year, and 0.915 over a 15-year time frame (Figure 2).

**Figure 2.**
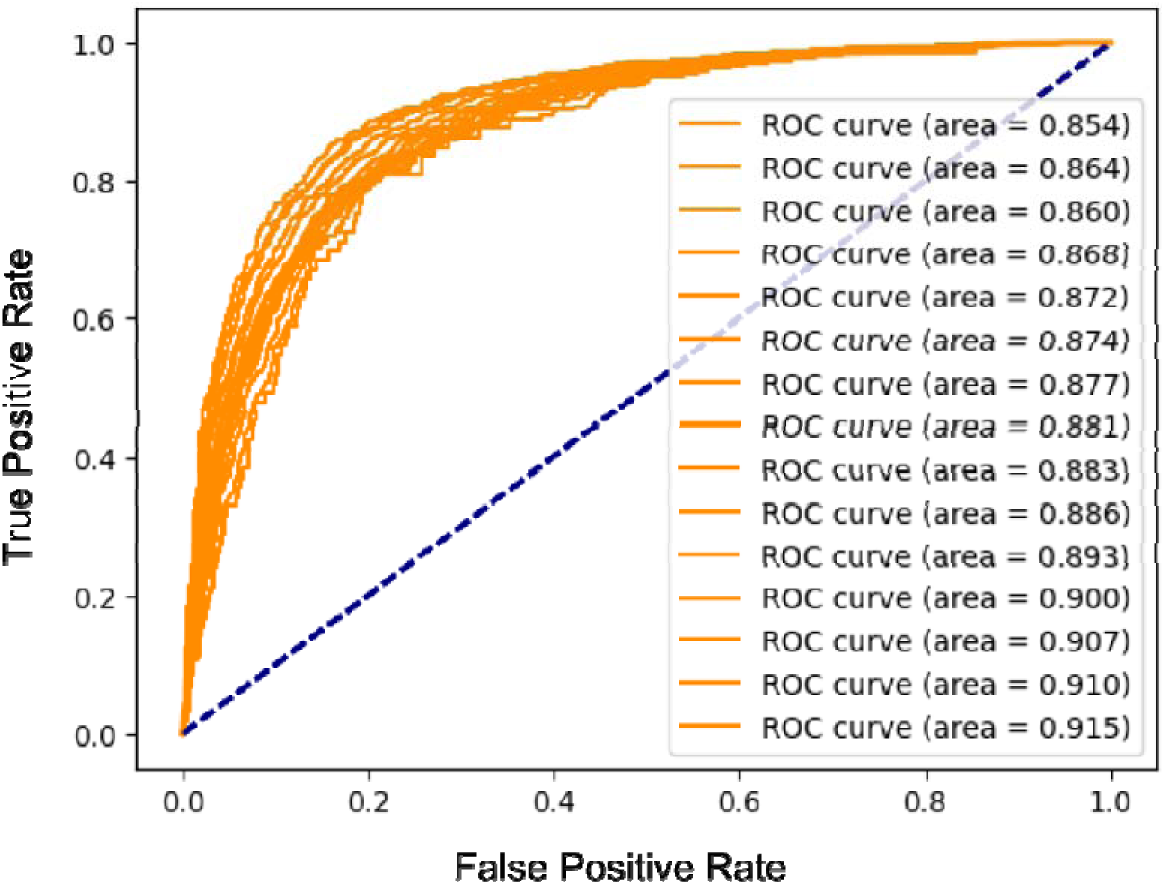
Performance of the Grocery Basket Score (GBS) Model. Areas under the receiver operating characteristic curve (ROC-AUCs) demonstrating the performance of the GBS Model when predicting mortality for NHANES participants on the test set over 1- to 15-year timeframes.

The dietary signals and respective regression coefficients of the model are presented in Table 1. The regression coefficients indicate that the model penalizes consuming calories from sugar, saturated fats, and beverages as well as high salt intake. Fiber consumption is rewarded. Furthermore, a penalty is applied to diets with a low ratio of protein, vitamin C, or iron consumption relative to the total calories.

**Table 1.** Dietary signals and regression coefficients of the core and beverage models of the Grocery Basket Score (GBS). Positive regression coefficients indicate increased mortality risk, while negative regression coefficients indicate decreased mortality risk. The low nutrient-energy-ratio signals were obtained by reviewing nutrition guidelines and dividing the daily recommended intake of the nutrient by the daily recommended calories from the same source, resulting in a recommended density of the nutrient per calorie. The low nutrient-energy-ratio signal is considered if the nutrient per calorie density in the basket is below the recommended density. The signal becomes stronger as the ratio becomes lower. To calculate the GBS for a person’s basket, each dietary signal is multiplied with the respective regression coefficient. The resulting output is the logarithm of the hazard ratio and undergoes the power shift described in the methodology section to create the final GBS on a scale of 0 to 1’000.

| Core model |  |  |  |
| --- | --- | --- | --- |
| Dietary signal | Regression coefficient | Mean prior to standardization | Standard deviation prior to standardization |
| Calories from saturated fats [kcal] / total calories [kcal] | 0.059 | 0.1208 | 0.0357 |
| Low protein-energy-ratio | 0.020 | 0.0001 | 0.0009 |
| Calories from sugar [kcal] / total calories [kcal] | 0.056 | 0.1662 | 0.0772 |
| Amount of fiber [g] / total calories [kcal] | - 0.081 | 0.090 | 0.0039 |
| Amount of sodium [mg] / total calories [kcal] | 0.064 | 1.8112 | 0.4730 |
| Low vitamin-C-energy-ratio | 0.153 | 0.0171 | 0.0147 |
| Low iron-energy-ratio | 0.084 | 0.0005 | 0.0011 |
| Beverage model |  |  |  |
| Dietary Signal | Regression coefficient | Mean prior to standardization | Standard deviation prior to standardization |
| Calories from beverages [kcal] / weight of beverages [g] | 0.071 | 0.1726 | 0.1451 |

As the direction of these coefficients was deemed to be in line with current nutritional research and the model offered the best compromise between predictive performance, comprehensiveness of the signals, and signal plausibility, it was selected as the first version of the Grocery Basket Score (GBS) model. Note that this selection represents a judgment call as selecting whichever combination of signals achieved the best performance could have yielded implausible coefficients due to multicollinearity.

A schema of performing inference with the GBS model is provided in Figure 3. In short, inference is performed by calculating the dietary signals, multiplying them with the respective regression coefficients, weighting core and beverage model by calorie contribution, and then performing the power shift as outlined in the methodology section.

**Figure 3.**
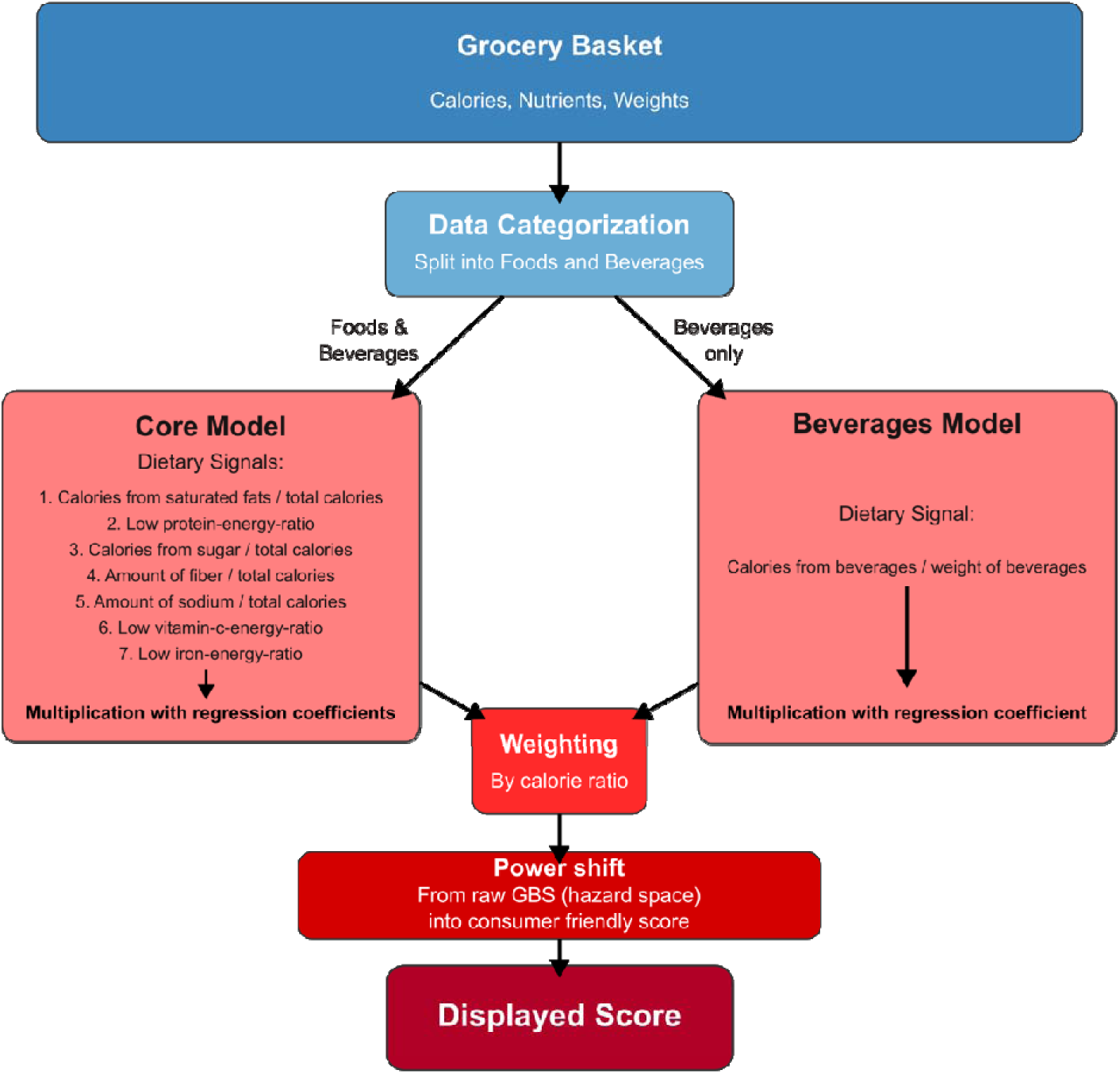
Schema of the Grocery Basket Score (GBS) model at inference. The calories, nutrients, and weights in the basket are summarized for foods and beverages respectively. The data for beverages are passed to the beverages model which only computes one dietary signal (calories from beverages / weight of beverages) and multiplies it with the respective regression coefficient. The data for both foods and beverages are passed to the core model, which computes seven dietary signals (calories from saturated fats / total calories, low protein-energy-ratio, calories from sugar / total calories, amount of fiber / total calories, amount of sodium / total calories, low vitamin-C-energy-ratio, low iron-energy-ratio) and multiplies them with the respective regression coefficients. The regression coefficients are presented in Table 1.

In addition, we have calculated the GBS for ten random NHANES participants using their dietary recalls with scores ranging from 262 to 709 and present this information in Table 2. The respective food items that the people consumed are provided in Supplementary Tables 4a and 4b.

**Table 2.**
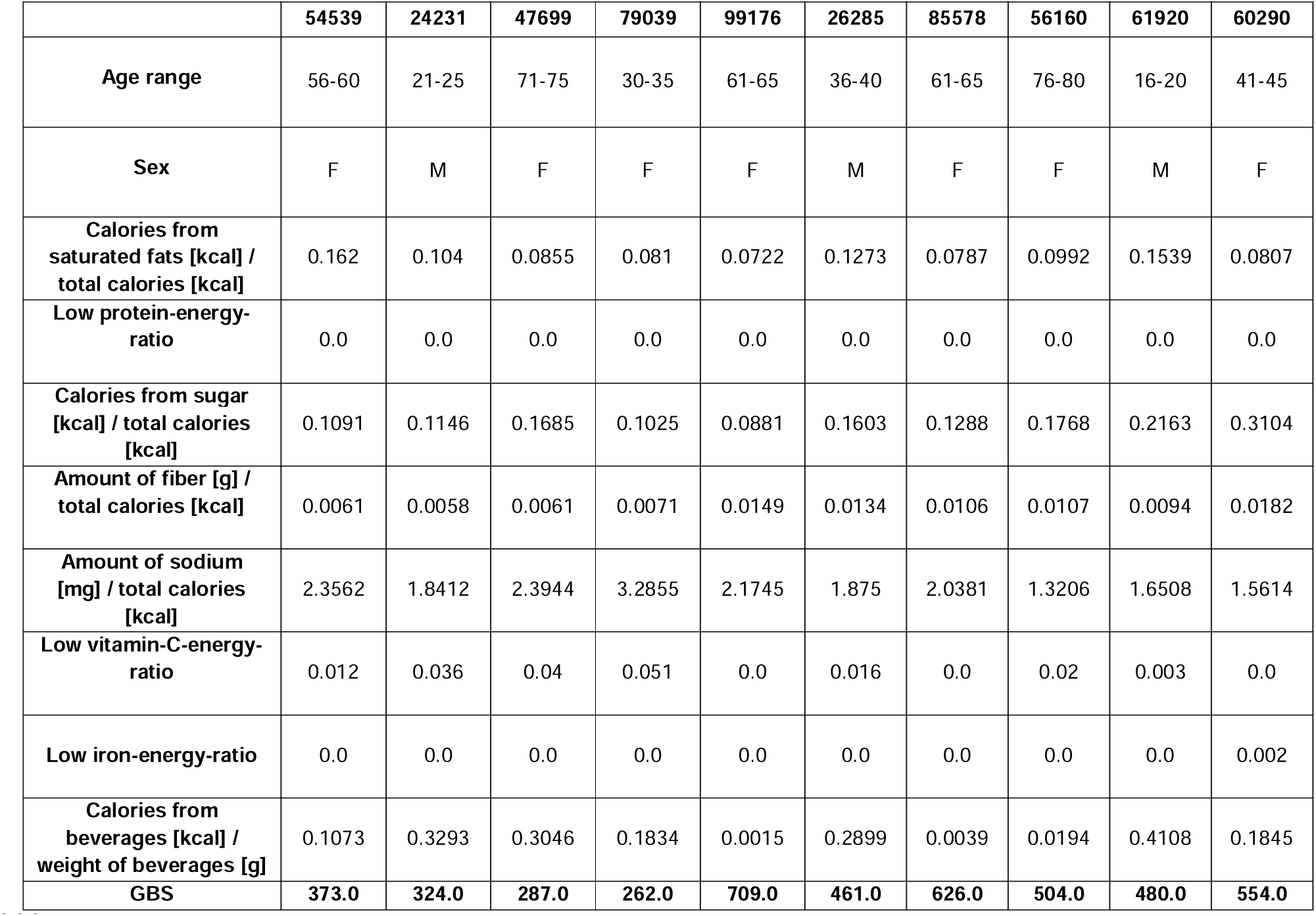
Grocery Basket Score (GBS) calculation for 10 random participants from NHANES using their answers to dietary recalls. The table presents the strength of the respective signals and the resulting scores. The consumed foods including the respective quantities are presented in Supplementary Table 4a (first five patients) and 4b (remaining patients). *F = female, M = male*.

To derive additional insights into possible correlations between signals, we also created a correlation matrix between different parameters based on items in the Open Food Facts database, which is presented in Figure 4.

**Figure 4.**
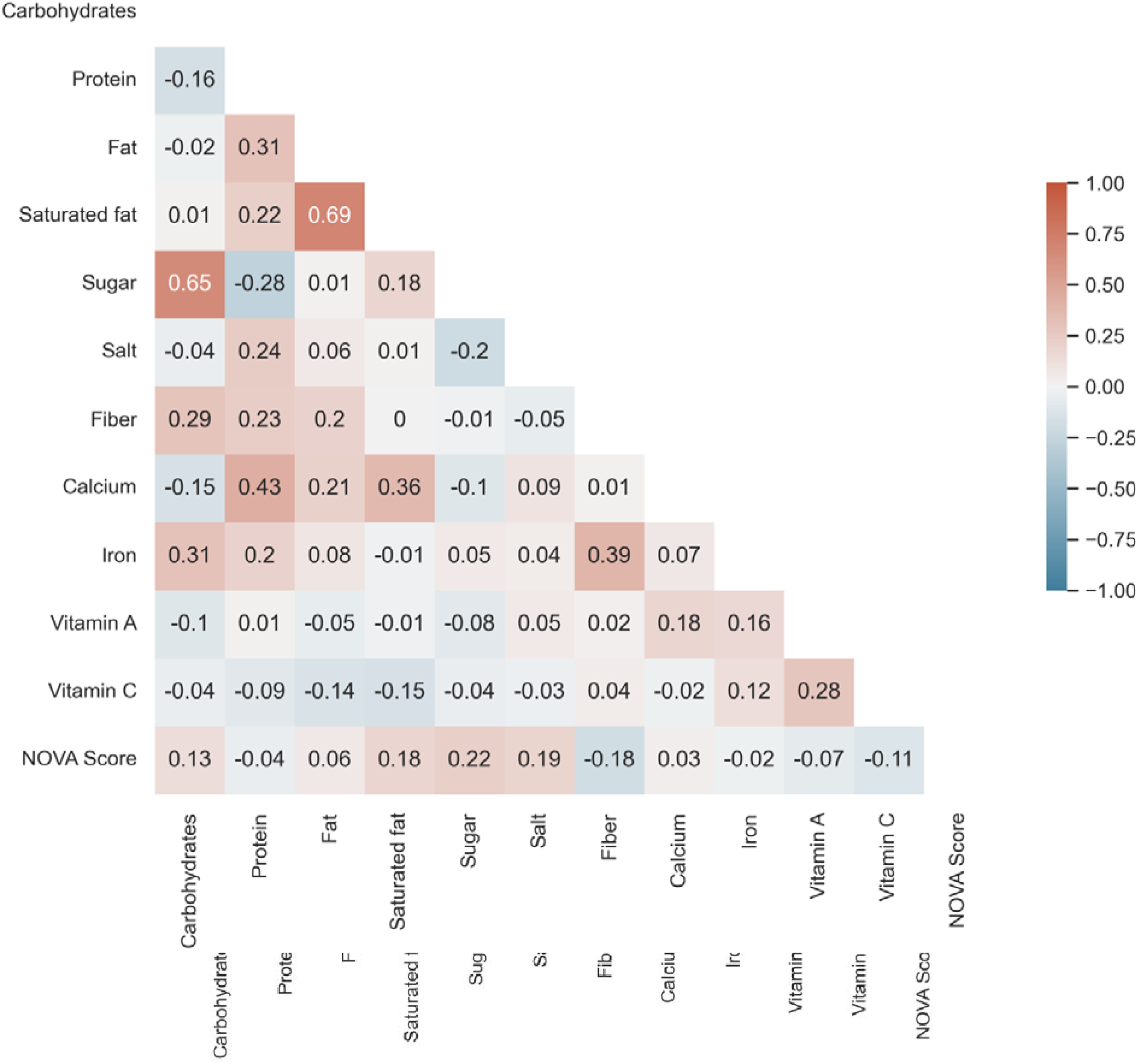
Pearson correlation coefficients between different parameters based on items in the Open Food Facts database. The NOVA Score tries to approximate the degree of processing a food item has undergone with higher scores indicating a higher degree.

### Validation

The correlation coefficients between the GBS and the AHEI for 4’382 participants in NHANES 2017-2018 were 0.62 for the AHEI without the protective effect of moderate alcohol consumption and 0.60 with it. The correlation coefficients between the calorie-weighted Nutri-Scores and the AHEI were -0.48 and -0.47, respectively. The corresponding scatterplots are presented in Figure 5.

**Figure 5.**
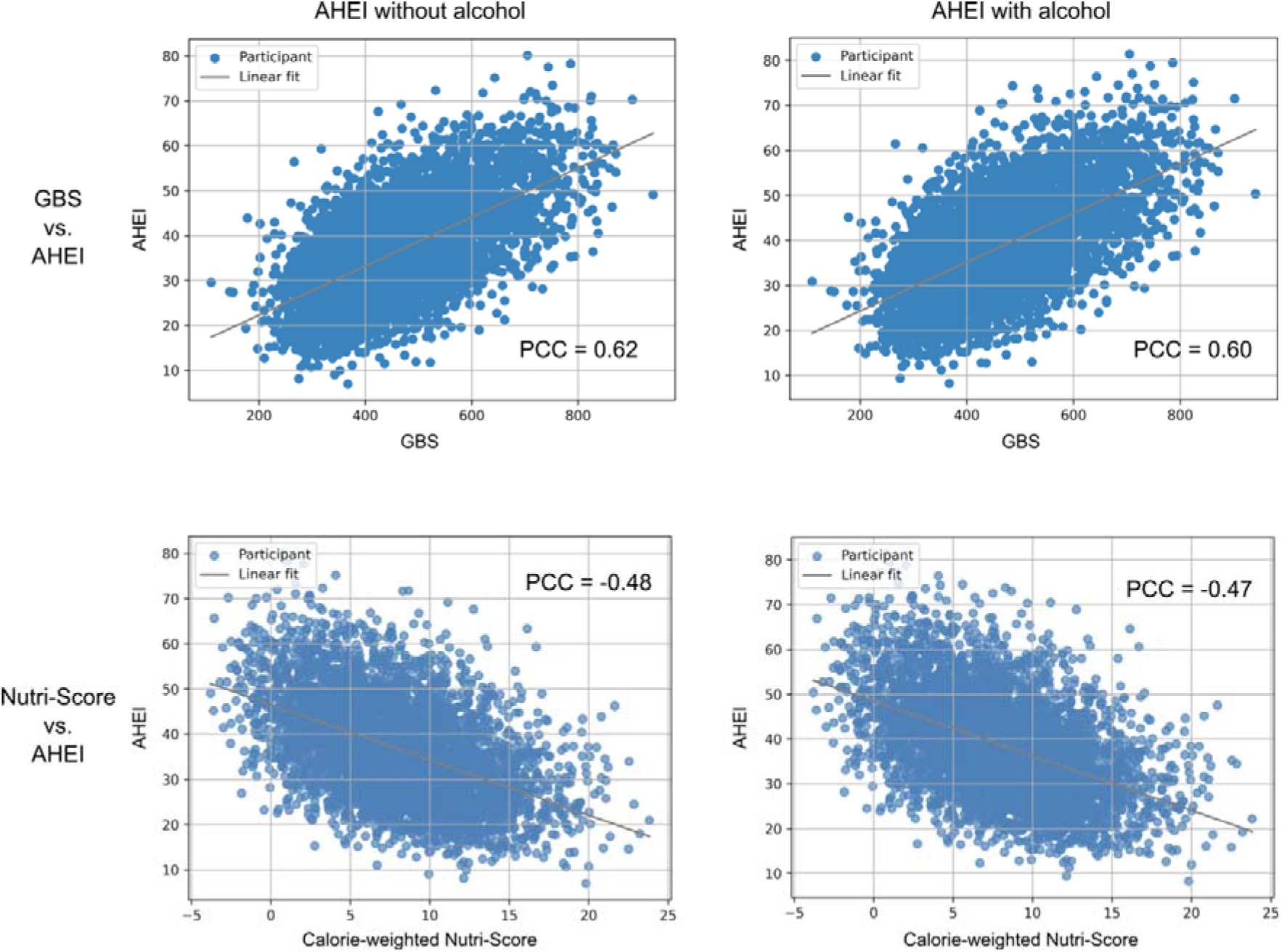
Scatterplot of the Grocery Basket Score (GBS - top) and calorie-weighted Nutri-Score (bottom) with Alternate Healthy Eating Index (AHEI) ratings. Each dot represents one of 4’382 NHANES 2017 - 2018 participants. Results of using the AHEI without the protective effect of moderate alcohol consumption are presented on the left and results with the protective effect of moderate alcohol consumption are displayed on the right. A steeper slope indicates a stronger correlation. *PCC = Pearson correlation coefficient*.

The overall correlation coefficient between the GBS and the Nutri-Score on single food items in the whole Open Food Facts database was -0.60. This inverse correlation is due to healthy items receiving low numeric values by the Nutri-Score but high numeric values by the GBS. The clustering of items from the Open Food Facts database resulted in 15 clusters, and the correlation coefficients per cluster ranged from -0.31 to -0.74. The clusters and respective correlation coefficients are provided in Table 3. The weakest correlation coefficient was found in cluster 7 with a correlation coefficient of -0.31, which was due to items being scored more favorably by the Nutri-Score than by the GBS. In all other clusters, correlation coefficients were stronger than -0.46.

**Table 3.** Pearson correlation coefficients by food clusters between the Grocery Basket Score (GBS) and the Nutri-Score. Greater absolute numbers indicate stronger correlations.

| Cluster # | Open Food Facts Tags | Pearson correlation coefficient |
| --- | --- | --- |
| 0 | seafood; fishes-and-their-products; fishes; fatty-fishes; canned-foods | -0.60 |
| 1 | fruits-and-vegetables-based-foods; plant-based-foods-and-beverages; plant-based-foods; vegetables-based-foods; canned-foods | -0.65 |
| 2 | biscuits-and-cakes; sweet-snacks; biscuits; snacks; cakes | -0.74 |
| 3 | meats-and-their-products; meats; prepared-meats; sausages; hams | -0.73 |
| 4 | dairies; fermented-foods; fermented-milk-products; cheeses; desserts | -0.46 |
| 5 | cereals-and-potatoes; plant-based-foods-and-beverages; plant-based-foods; cereals-and-their-products; breads | -0.68 |
| 6 | condiments; sauces; groceries; plant-based-foods-and-beverages | -0.59 |
| 7 | salty-snacks; appetizers; milks; chips-and-fries; crisps | -0.31 |
| 8 | meals; pasta-dishes; pizzas-pies-and-quiches; soups; meals-with-meat | -0.53 |
| 9 | frozen-foods; desserts; frozen-desserts; ice-creams-and-sorbets; dairy-desserts | -0.58 |
| 10 | confectioneries; sweet-snacks; snacks; cocoa-and-its-products; chocolate-candies | -0.53 |
| 11 | snacks; sweet-snacks; confectioneries; biscuits-and-cakes; biscuits | -0.74 |
| 12 | spreads; breakfasts; sweet-spreads; plant-based-spreads; fruit-and-vegetable-preserves | -0.66 |
| 13 | fats; vegetable-fats; vegetable-oils; plant-based-foods-and-beverages; plant-based-foods | -0.72 |
| 14 | beverages | -0.60 |
| **Overall** |  | **-0.60** |

Figure 6 shows the percentage of items per food cluster where the z-scores between the GBS and the Nutri-Score differ by more than 1 and 2 as well as the direction of the disagreement (i.e., which model rated more favorably). Samples of items scored differently by the GBS compared to the Nutri-Score are provided in Supplementary Tables 5 - 8.

**Figure 6.**
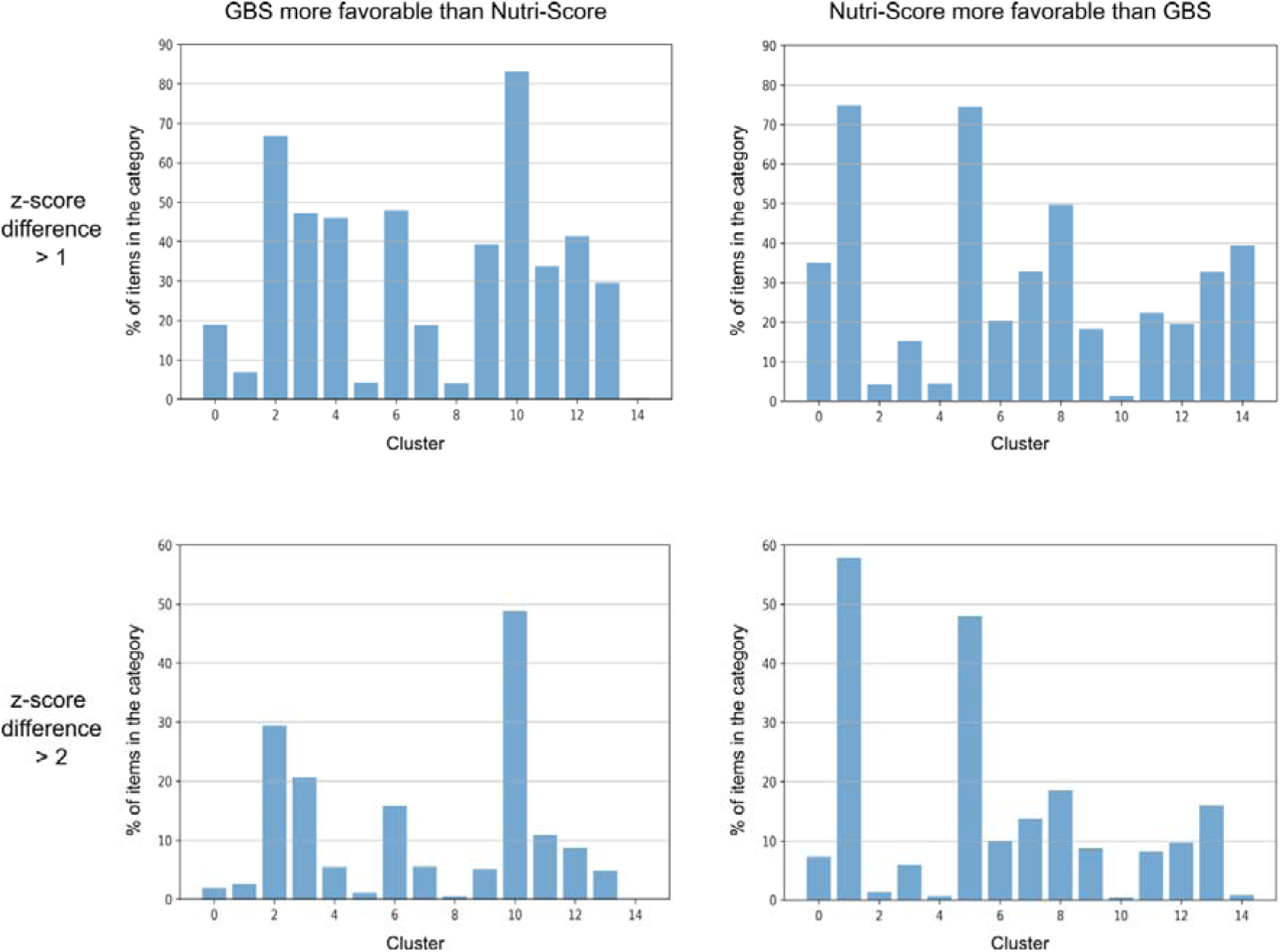
Discrepancies in Grocery Basket Score (GBS) and Nutri-Score ratings by food clusters. Percentage of items per food cluster where the z-scores between the GBS and the Nutri-Score differ by more than 1 (top) and 2 (bottom). Items that were rated more favorably by the GBS than the Nutri-Score are presented on the left, while items rated more favorably by Nutri-Score can be found on the right. The contents of the clusters can be found in Table 3.

The outputs of the core and beverage models are weighted by the ratio of the total calories handled by the respective models. If a basket’s calories come almost exclusively from food, the output of the core model will dictate the total score. If a basket’s calories come almost exclusively from beverages, the output of the beverage model will dictate the total score. The power shift from the resulting logarithm of the hazard ratio to the 0 to 1’000 scale is performed according to the description in the methods section.

## Discussion

The main findings of this study are that the novel nutrient energy density model tested, predicted healthy diet scores that, like Nutri-score and AHEI, are in alignment with the nutrition guidelines issued by the U.S. Departments of Agriculture and of Health and Human Services or the World Health Organization (13,14). This new nutrient energy density model therefore can now be used as a Grocery Basket Score (GBS) to automatically score foods and beverages in grocery shopping baskets according to their health properties based on their link to mortality. Its final score is computed as a calorie-weighted average of two separate models, one scoring a combination of foods and beverages and the other once scoring only beverages.

The core model’s characteristics can be derived from the regression coefficients assigned to its nutrient energy density signals: It penalizes consuming calories from sugar and saturated fats, as well as high salt intake. Furthermore, a penalty is applied to baskets with a low ratio of protein, vitamin C, or iron, relative to their total calories. Fiber consumption is rewarded. For the beverages model, calorie density is penalized.

Even though the GBS was designed to be deployed on whole shopping baskets and not for assessing individual products, the high degree of correlation with the Nutri-Score, indicated by a correlation coefficient of -0.60 on items from the Open Food Facts database, demonstrates the considerable overlap in principles such as rewarding the presence of fiber and penalizing the presence of saturated fats, sugar, and salt. This should reduce the risk of confusing consumers should the GBS be deployed in regions where the Nutri-Score is also used for individual products.

The design differences to the Nutri-Score become evident when looking at the food clusters with lower correlation coefficients and the items for which the scores disagreed the most. The differences in the scoring of items in the cluster with the lowest absolute correlation coefficient (i.e., cluster 7) appear to be mainly driven by (low-fat) milk, which is rated more negatively by the GBS than by the Nutri-Score. The same applies to cluster 4 where the differences appear to be driven by dairy products such as cottage cheese or yoghurt. This is due to the lack of vitamin C, iron, and fiber, which is penalized by our model as a balanced shopping basket should contain these nutrients. However, milk and egg whites are obviously not supposed to be sources of fiber. When part of a more comprehensive shopping basket, they would have a more positive effect due to improving a low protein-energy-ratio and increasing the denominator (i.e., the total calories), thereby improving the sugar and saturated fat divided by total calorie signals.

Cluster 10 contains sweet snacks, such as chocolate, which tend to be scored more positively by the GBS than by the Nutri-Score. This seems to be mainly because the Nutri-Score considers calorie density as a standalone signal and punishes items for having high calorie densities. The GBS takes a different route as it has been adjusted for BMI and does not consider calorie density as its own signal, effectively asking the question “If a consumer is able to maintain their weight, is it ok for them to consume this shopping basket?”. While weight management is certainly an important goal when trying to compose a shopping basket, it is also relatively easy to track for the consumer when stepping on a scale. The GBS thus focuses on issues that are more difficult to keep track of, such as the nutrient composition, which indirectly relates to weight management as fiber and protein tend to provide more satiety per calorie than sugars.

The high degree of correlation with the AHEI is promising and not surprising considering that the AHEI advises people to frequently eat fiber- and protein-rich foods while lowering sugar and salt consumption. It is also unsurprising that the correlation is even better when using the AHEI without assuming a protective effect of moderate alcohol consumption as the beverage model penalizes drinks with higher calorie densities.

Our study has several limitations. Firstly, our grocery basket was based on all foods consumed over 24h, as recalled by participants in the NHANES study. It represented one person’s daily diet. Although isolated grocery baskets in real life are unlikely to contain all of one person’s daily diet, over a period of time, combined grocery baskets can be representative of a single person’s or whole family’s diet. Then, the greater number of GB scores from a single or multiple supermarkets will improve the overall evaluation of the health value of a family diet and the consumer can also be advised on how to improve the diet.

A general limitation of working with the NHANES dataset is its cross-sectional nature. That is, a person’s nutrition might have changed significantly from the time of their dietary recalls to their death or time of censoring. Furthermore, the fact that some signals are included in the final model while others are not, should not be interpreted as a claim that the included signals are somehow more predictive of mortality, let alone that there is more evidence for a causal relation, i.e., that supplementing the respective nutrients will improve mortality. It seems plausible that some of the signals used in the GBS function largely as proxies, such as the fiber signal and the vitamin-C-energy-ratio signal being proxies for fruit and vegetable or whole grains consumption which need to be considered when using the GBS to propose a healthy diet.

Furthermore, it would have been desirable to have even more signals to get a more comprehensive picture of someone’s nutrition. However, due to the high degree of correlation between many signals, including more signals would not only provide, at best, minimal lift but also make the modeling more unstable due to variables stealing predictive power from each other (15). While there are certainly other ways to do the modeling, and judgment calls of some degree were necessary to make the signal selection, we believe that the current model is capable of giving the consumer a useful evaluation of the quality of their diet, especially if grocery basket scores from a single retailer, or groups of retailers, are combined over a period of time.

The GBS is also relatively resistant to current attempts at gaming the system, e.g., by adding soluble fiber or proteins to beverages as this would not improve the beverage model that only focuses on calorie density. While having the degree of processing as a signal would have been interesting, this is harder to do without requiring manual labeling. In addition, ultra-processed foods are frequently high in salt, sugar, and saturated fats but relatively low in protein and fiber, as illustrated by the correlation matrix in Figure 4, so the degree of processing is at least indirectly represented in the GBS through surrogate signals (16). This is supported by a recent study by Fang and colleagues, who found that adjusting for the AHEI attenuated a previously observed association of ultra-processed food intake with mortality (17). The annotation problem and the representation through surrogates also apply to other potential signals like including whole grains, which have been suggested previously (18).

Other limitations include the fact that this model was developed on a U.S. instead of a global population which might limit its ability to generalize to other parts of the world. In addition, the dietary information was self-reported. Furthermore, food items with missing nutritional information were not considered during the validation, which might have introduced bias, as well as the fact that for technical reasons Nutri-Score 2017 was used for the AHEI validation instead of the more recent version 2023.

Lastly, another limitation of the GBS is that it does not model the negative health effects of alcoholic beverages that go beyond their calorie density (19). The fact that adding the self-reported alcohol consumption as a signal did not increase the performance of the model when predicting mortality in NHANES might be due to inconsistencies in self-reported and actual alcohol consumption that have previously been described (20).

A strength of the GBS is that it does not require manual annotation of food items prior to deployment, as it leverages the Open Food Facts database and an imputation engine. Since many retailers already have loyalty or digital shopping basket programs in place, this allows for automated real-world implementation. The use of high-level signals yielding a fingerprint, as opposed to very granular lists of food items people should consume, is aligned with the quality of evidence in the nutritional epidemiology space and allows for flexibility when composing a basket.

Most importantly, the GBS gives users an automated assessment of their entire shopping basket without the need for manual tracking. Moreover, owing to its energy density-based nature, GBS does not require absolute quantities or knowledge of the household size. Even with shopping at multiple grocers and eating out, GBS can provide a reasonable score as long as a representative subset of food intake is scored. As an outlook, the model could also be used to derive personal shopping recommendations based on the strengths of the signals for a given basket.

In conclusion, the Grocery Basket Score (GBS) based on a nutrient energy density model is a methodology for scoring shopping baskets that goes beyond averaging the scores of individual products. The GBS scoring methodology is aligned with the recommendations formulated in nutritional guidelines, which is indicated by a high degree of correlation with the AHEI. Product-level profiling systems such as the Nutri-Score also exhibit a correlation with the GBS and can be used in combination.

## Disclosures

P.W., S.M., J.O., D.F., R.H., and A.N. and. report employment at or consulting for dacadoo AG, a technology company that has business activity with retailers, including grocery stores. dacadoo intends to file ‘GBS’ as a trademark and commercialize GBS^TM^ software and/or consulting services. The other authors declare no additional conflict of interest.

## Author contributions

Conceptualization, P.W., A.N.; methodology, P.W, S.M., A.N.; formal analysis, P.W., S.M., J.O., A.N.; data curation, S.M., J.O.; writing—original draft preparation, P.W.; writing—review and editing, S.M., J.O., D.F., R.H., A.N.; supervision, A.N.; project administration, P.W., A.N.;

## Data availability

This paper contains information from the Open Food Facts (https://world.openfoodfacts.org/) and OFF-GBS (https://github.com/windisch-paul/OFF-GBS) databases, which are made available under the Open Database License (ODbL, https://opendatacommons.org/licenses/odbl/1-0/).

## Ethical approval

Not applicable (publicly available, anonymized data)

## Funding

No external funding was received for this project.

## Notes

### Author Declarations

The study used openly available human data from NHANES (https://www.cdc.gov/nchs/nhanes/index.html)

### Summary of Updates

We have appended the manuscript with additional figures, tables, and explanations to make it more understandable to a non-data science audience. Two researchers who were involved in this have been added as co-authors.

