## Supplementary tables 1, 2, 3. for "Development and Validation of a Nutrient Profiling Model for Shopping Baskets: The Grocery Basket Score (GBS) Methodology"

| Nutrient | RDA | Recommended calories per day |
|---|---|---|
| Vitamin C | M: 90 mg<br>F: 75 mg<br><br>+ 35 mg for smokers | |
| Folate | M: 400 µg | |
| Potassium | M: 3400 mg<br>F: 2600 mg | M: 2’500 kcal<br>F: 2’000 kcal |
| Calcium | < 50 y/o: 1’000 mg<br>>= 50 y/o: 1’200 mg | |
| Protein | M: 60 g<br>F: 48 g | |
| Iron | M, F> 50: 8 mg<br>F =< 50: 18 mg | |

**Supplementary Table 1. Nutrient and calorie recommendations that were used as the basis for low nutrient-energy-ratio signals.** *RDA = Recommended daily allowance; M = male; F = female.*

| Variable | Adjusted by age, sex | Adjusted by age, sex, BMI | Adjusted by age, sex, BMI, PIR |
|---|---|---|---|
| **Age** | 1.882 | 1.884 | 1.831 |
| **Sex** | 0.169 | 0.169 | 0.189 |
| **BMI** | | 0.011 | 0.008 |
| **PIR** | | | -0.251 |
| **Calories from saturated fats / total calories** | 0.046 | 0.045 | 0.059 |
| **Protein deficiency** | 0.023 | 0.023 | 0.020 |
| **Calories from sugar / total calories** | 0.038 | 0.038 | 0.056 |
| **Amount of fiber / total calories** | -0.081 | -0.081 | -0.081 |
| **Amount of sodium / total calories** | 0.064 | 0.064 | 0.064 |
| **Vitamin C deficiency** | 0.192 | 0.192 | 0.153 |
| **Iron deficiency** | 0.105 | 0.106 | 0.084 |

**Supplementary Table 2. Adjustments of the core model and respective coefficients.** *BMI = Body mass index; PIR = poverty income ratio.*

| Variable | Adjusted by age, sex | Adjusted by age, sex, BMI | Adjusted by age, sex, BMI, PIR |
|---|---|---|---|
| **Age** | 1.786 | 1.789 | 1.755 |
| **Sex** | 0.168 | 0.169 | 0.190 |
| **BMI** | | 0.016 | 0.010 |
| **PIR** | | | -0.278 |
| **Calories from beverages / weight of beverages** | 0.078 | 0.079 | 0.071 |

**Supplementary Table 3. Adjustments of the beverages model and respective coefficients.** *BMI = Body mass index; PIR = poverty income ratio.*

**Supplementary Table 4a. Sample Grocery Basket Scores a.** Grocery Basket Score (GBS) calculation for 5 random participants from NHANES using their answers to dietary recalls. See the attached file “supplementary_table_8a.pdf”.

**Supplementary Table 4b. Sample Grocery Basket Scores b.** Grocery Basket Score (GBS) calculation for an additional 5 random participants from NHANES using their answers to dietary recalls. See the attached file “supplementary_table_8b.pdf”.

**Supplementary Table 5. 250 random items from the GBS x Nutri-Score validation where the difference of the z-scores was > 1, and the GBS scored the items more favorably than the Nutri-Score.** See the attached file “supplementary_table_4.csv”.

**Supplementary Table 6. 250 random items from the GBS x Nutri-Score validation where the difference of the z-scores was > 1, and the GBS scored the items more negatively than the Nutri-Score.** See the attached file “supplementary_table_5.csv”.

**Supplementary Table 7. 250 random items from the GBS x Nutri-Score validation where the difference of the z-scores was > 2, and the GBS scored the items more favorably than the Nutri-Score.** See the attached file “supplementary_table_6.csv”.

**Supplementary Table 8. 250 random items from the GBS x Nutri-Score validation where the difference of the z-scores was > 2, and the GBS scored the items more negatively than the Nutri-Score.** See the attached file “supplementary_table_7.csv”.
