## Supplementary table 4a for "Development and Validation of a Nutrient Profiling Model for Shopping Baskets: The Grocery Basket Score (GBS) Methodology"

| | 54539 | 24231 | 47699 | 79039 | 99176 |
|---|---|---|---|---|---|
| **age** | 56-60 | 21-25 | 71-75 | 31-35 | 61-65 |
| **sex** | F | M | F | F | F |
| **Calories from saturated fats [kcal] / total calories [kcal]** | 0.162 | 0.104 | 0.0855 | 0.081 | 0.0722 |
| **Low protein-energy-ratio** | 0.0 | 0.0 | 0.0 | 0.0 | 0.0 |
| **Calories from sugar [kcal] / total calories [kcal]** | 0.1091 | 0.1146 | 0.1685 | 0.1025 | 0.0881 |
| **Amount of fiber [g] / total calories [kcal]** | 0.0061 | 0.0058 | 0.0061 | 0.0071 | 0.0149 |
| **Amount of sodium [mg] / total calories [kcal]** | 2.3562 | 1.8412 | 2.3944 | 3.2855 | 2.1745 |
| **Low vitamin-C-energy-ratio** | 0.012 | 0.036 | 0.04 | 0.051 | 0.0 |
| **Low iron-energy-ratio** | 0.0 | 0.0 | 0.0 | 0.0 | 0.0 |
| **Calories from beverages [kcal] / weight of beverages [g]** | 0.1073 | 0.3293 | 0.3046 | 0.1834 | 0.0015 |
| **GBS** | 373.0 | 324.0 | 287.0 | 262.0 | 709.0 |
| **Liquids** | COFFEE, MADE FROM GROUND, REGULAR: 266.4g<br>MILK, COW'S, FLUID, 2% FAT: 61.0g<br>ORANGE JUICE, W/ CALCIUM, CAN/BOTTLED/CARTON: 155.0g<br>SOFT DRINK, COLA-TYPE, SUGAR-FREE: 121.5g<br>TEA, LEAF, UNSWEETENED: 177.6g<br>MILK, COW'S, FLUID, 2% FAT: | MILK, COW'S, FLUID, 2% FAT: 183.0g<br>FRUIT-FLAVORED DRINK, LOW CAL, W/ VITAMIN C ADDED: 525.0g<br>COLA W/ FRUIT OR VANILLA FLAVOR: 806.0g<br>MILK, COW'S, FLUID, 2% FAT: 122.0g<br>SOFT DRINK, COLA-TYPE: 992.0g | WATER, TAP: 474.0g<br>FRUIT JUICE DRINK: 124.0g<br>MILK, COW'S, FLUID, 1% FAT: 244.0g<br>NONALCOHOLIC MALT BEVERAGE: 360.0g<br>SOFT DRINK, FRUIT-FLAVORED, CAFFEINE FREE: 246.4g<br>WATER AS AN INGREDIENT: 44.44g<br>SOFT DRINK, COLA-TYPE, | SOFT DRINK, FRUIT-FLAVORED, W/ CAFFEINE: 372.0g<br>WATER, TAP: 2100.0g<br>SOFT DRINK, FRUIT-FLAVORED, W/ CAFFEINE: 372.0g<br>TEA, ICED, BREWED, GREEN, PRE-SWEETENED WITH SUGAR: 241.5g<br>SOFT DRINK, FRUIT-FLAVORED, W/ CAFFEINE: | COFFEE, BREWED: 150.0g<br>MILK, LOW FAT (1%): 30.5g<br>WATER, BOTTLED, UNSWEETENED: 507.0g<br>WATER, BOTTLED, UNSWEETENED: 507.0g<br>WATER, BOTTLED, UNSWEETENED: 360.0g<br>COFFEE, BREWED: 240.0g<br>WATER, BOTTLED, UNSWEETENED: 120.0g<br>WATER, BOTTLED, |

| | | | | | |
|---|---|---|---|---|---|
| | 30.5g<br>TEA, LEAF, UNSWEETENED: 518.0g<br>MILK, COW'S, FLUID, 2% FAT: 61.0g<br>PRUNE JUICE: 160.0g<br>COFFEE, MADE FROM GROUND, REGULAR: 310.8g<br>CREAM, HALF & HALF: 30.25g<br>COFFEE, MADE FROM GROUND, REGULAR: 177.6g<br>CREAM, HALF & HALF: 37.81g<br>WATER, BOTTLED, UNSWEETENED: 14.81g | | DECAFFEINATED: 368.4g<br>ORANGE JUICE, CANNED, BOTTLED OR IN A CARTON: 210.0g | 372.0g<br>SOFT DRINK, FRUIT-FLAVORED, W/ CAFFEINE: 186.0g<br>SOFT DRINK, FRUIT-FLAVORED, W/ CAFFEINE: 248.0g<br>WATER, BOTTLED, UNSWEETENED: 507.0g<br>SOFT DRINK, FRUIT-FLAVORED, W/ CAFFEINE: 372.0g<br>TEA, ICED, BREWED, BLACK, DECAF, PRE-SWEETENED W/SUGAR: 372.0g<br>WATER, BOTTLED, UNSWEETENED: 507.0g<br>WATER, BOTTLED, UNSWEETENED: 507.0g<br>MILK, WHOLE: 76.25g | UNSWEETENED: 507.0g<br>WATER, BOTTLED, UNSWEETENED: 253.5g |
| **Meat-based dishes** | HAM, SLICED, LOW SALT, PREPACKAGED/DELI, LUNCH MEAT: 14.0g<br>CHALUPA W/ BEEF, CHEESE, LETTUCE, TOMATO & SOUR CREAM: 156.0g<br>BEEF & VEG (W/ CAR/DK GREEN, NO POTATO), SOY SAUCE: 284.81g<br>CHICKEN, NS AS TO PART, NS METHOD, SKIN: 67.5g<br>LIVERWURST: 106.4g | DOUBLE BACON CHEESEBURGER, ON BUN: 290.0g<br>CHICKEN, BREAST, ROASTED/BROILED/BAKED, NS SKIN: 226.8g<br>MEAT LOAF W/ BEEF, W/ TOMATO SAUCE: 183.0g<br>MEATLOAF, NS AS TO TYPE OF MEAT: 63.79g | CHICKEN,WING,COATED,BKD/FRD,PPD W/SKIN, SKIN EATEN: 84.0g<br>CHICKEN, WING, ROASTED/BROILED/BAKED, W/O SKIN: 24.0g<br>BACON, NS AS TO TYPE OF MEAT, COOKED: 0.63g | HAM & CHEESE SUB, W/ LETTUCE, TOMATO & SPREAD: 468.0g<br>HAM, SLICED, PREPACKAGED OR DELI, LUNCHEON MEAT: 56.0g<br>HAM, SLICED, PREPACKAGED OR DELI, LUNCHEON MEAT: 75.6g<br>BACON, NS AS TO TYPE OF MEAT, COOKED: 2.5g | CHICKEN FILLET SANDWICH, GRILLED, FROM FAST FOOD: 70.0g<br>KUNG PAO CHICKEN: 91.13g<br>CHICKEN THIGH, ROTISSERIE, SKIN NOT EATEN: 170.1g<br>PORK CHOP, BROILED OR BAKED, LEAN ONLY EATEN: 84.0g |
| **Eggs, Cheese, and Yoghurt** | EGGS, WHOLE, FRIED (INCL SCRAMBLED, NO MILK ADDED): 46.0g<br>CHEESE, MONTEREY: 42.38g<br>CHEESE,CHEDDAR/AMERICAN TYPE,NS NATURAL OR PROCESSED: 56.5g | nan | POTATO SALAD W/ EGG: 36.19g | CHEESE, AMERICAN: 21.0g<br>CHEESE, SWISS: 14.0g | EGG, WHOLE, BOILED OR POACHED: 50.0g<br>EGG, WHOLE, BOILED OR POACHED: 25.0g |

| | | | | | |
|---|---|---|---|---|---|
| **Grains, Rice, and Noodles** | ROLL, HOAGIE, SUBMARINE,: 36.5g<br>RICE, FRIED, W/ PORK: 148.5g<br>NOODLES, CHOW MEIN: 25.31g<br>TORTILLA, FLOUR (WHEAT): 67.5g<br>CORN FLAKES, KELLOGG'S: 21.0g<br>CRACKERS, SALTINES: 6.0g<br>BREAD, RYE: 60.8g | FROSTED FLAKES, KELLOGG: 89.69g<br>BREAD, WHITE: 40.0g<br>SPECIAL K CEREAL: 31.0g<br>SPAGHETTI, COOKED, NO FAT ADDED: 480.0g<br>BREAD, GARLIC: 153.6g | RICE KRISPIES, KELLOGG'S: 22.75g<br>BISCUIT, BAKING POWDER/BUTTERMILK TYPE, COMMERCIALLY BAKED: 57.0g<br>CRACKER, SNACK: 12.8g | BREAD, WHITE: 56.0g<br>ROLL, WHITE, HOAGIE, SUBMARINE: 48.67g<br>RICE, WHITE, COOKED, FAT NOT ADDED IN COOKING: 79.0g<br>CAP'N CRUNCH CEREAL: 27.0g | BREAD, FRENCH OR VIENNA: 50.66g<br>RICE, BROWN, COOKED, NS AS TO FAT: 257.25g<br>RICE, BROWN, COOKED, NS AS TO FAT: 196.0g<br>RICE, BROWN, COOKED, NO ADDED FAT: 195.0g<br>PHO: 533.75g |
| **Snacks** | LIGHT ICE CREAM, NO SUGAR ADDED, NOT CHOCOLATE: 129.5g | ALMONDS, SUGAR-COATED (INCL JORDAN ALMONDS): 17.5g | WHITE POTATO, CHIPS: 14.0g<br>CAKE, YELLOW, W/ ICING, HOMEMADE: 54.4g | nan | nan |
| **Fruits** | PINEAPPLE, COOKED OR CANNED, DRAINED SOLIDS: 179.0g<br>BANANA, RAW: 50.5g | nan | nan | nan | BANANA, RAW: 126.0g<br>BANANA, RAW: 126.0g |
| **Vegetables, Beans, and Lentils** | WHITE POTATO, HASH BROWN, FROM FROZEN: 72.5g<br>LETTUCE, RAW: 27.5g<br>TOMATOES, RAW: 135.0g<br>PEPPERS, HOT, COOKED, NS FORM, NS FAT ADDED: 2.2g<br>CUCUMBER PICKLES, DILL: 35.0g<br>LETTUCE, RAW: 55.0g<br>TOMATOES, RAW: 51.0g<br>ONIONS, MATURE, RAW: 4.5g<br>CUCUMBER, RAW (INCLUDE CUCUMBER, NFS): 21.0g | WHITE POTATO, FRENCH FRIES, FROM FROZEN, DEEP-FRIED: 63.75g<br>ENDIVE, CHICORY, ESCAROLE OR ROMAINE LETTUCE, RAW: 42.53g | VEGETABLE SOUP, CHUNKY STYLE: 170.1g<br>VEGETABLE SOUP, CANNED, LOW SODIUM: 340.2g<br>BEANS, STRING, CKD, FROM CAN, NS COLOR, FAT ADDED: 17.5g | WHITE POTATO, STICKS: 0.3g<br>CORN, YELLOW, COOKED, FROM CANNED, FAT NOT ADDED: 41.0g<br>BEANS, STRING, GREEN, COOKED, FROM CANNED, FAT NOT ADDED: 76.5g | LETTUCE, FOR USE ON A SANDWICH: 4.0g<br>MUSHROOMS, FOR USE ON A SANDWICH: 6.0g<br>ONIONS, FOR USE ON A SANDWICH: 7.5g<br>TOMATOES, FOR USE ON A SANDWICH: 10.0g<br>CARROTS, RAW: 40.0g<br>ROMAINE LETTUCE, RAW: 26.0g<br>BEAN SPROUTS, RAW: 90.0g<br>ONIONS, GREEN, RAW: 80.0g<br>AVOCADO, RAW: 150.0g<br>BROCCOLI, COOKED, FROM RESTAURANT: 160.0g<br>BROCCOLI, CHINESE, COOKED: 113.4g<br>GREEN BEANS, FRESH, |

| | | | | | |
|---|---|---|---|---|---|
| | | | | | COOKED, NO ADDED FAT: 56.7g |
| **Nuts** | PEANUT BUTTER: 16.0g | nan | nan | nan | MIXED NUTS, WITHOUT PEANUTS, UNSALTED: 71.0g |
| **Other** | SUCRALOSE-BASED SWEETENER, SUGAR SUBSTITUTE: 2.0g<br>SALSA, RED, CKD, NOT HOM (INCL TACO, CREOLE, PICANTE SAUCES): 5.19g<br>SOY SAUCE: 10.63g<br>WON TON (WONTON) SOUP: 527.19g<br>EGG ROLL, MEATLESS: 26.0g<br>DUCK SAUCE (INCLUDE CHAISNI SAUCE): 96.66g<br>BUTTER-MARGARINE BLEND, STICK, SALTED: 9.46g<br>SUGAR SUBSTITUTE, ASPARTAME-BASED, DRY POWDER: 2.0g<br>SUGAR SUBSTITUTE, SACCHARIN-BASED, DRY POWDER AND TABLETS: 2.0g<br>SUGAR SUBSTITUTE, SACCHARIN-BASED, DRY POWDER AND TABLETS: 1.0g<br>SUGAR SUBSTITUTE, ASPARTAME-BASED, DRY POWDER: 1.0g<br>MUSTARD (INCL HORSERADISH MUSTARD, CHINESE MUSTARD): 1.19g<br>ITALIAN DRESSING, LOW CALORIE: 30.0g<br>SUGAR SUBSTITUTE, SACCHARIN-BASED, DRY POWDER AND TABLETS: 2.0g | SUGAR, WHITE, GRANULATED OR LUMP: 25.0g<br>CAESAR DRESSING: 14.69g<br>SPAGHETTI SAUCE, MEATLESS: 170.1g | nan | SALAD DRESSING, NFS, FOR SANDWICHES: 19.07g<br>GRAVY, BEEF/MEAT (INCL GRAVY,NFS;BROWN GRAVY;SWISS STEAK GRV: 43.69g | SOY BASED SAUCE, FOR USE WITH VEGETABLES: 60.0g<br>COFFEE CREAMER,POWDER, SUGAR FREE, FLAVORED: 5.88g<br>SUGAR SUBSTITUTE, SUCRALOSE, POWDER: 0.5g<br>SOY BASED SAUCE, FOR USE WITH VEGETABLES: 77.4g |
