## Supplementary table 4b for "Development and Validation of a Nutrient Profiling Model for Shopping Baskets: The Grocery Basket Score (GBS) Methodology"

| | 26285 | 85578 | 56160 | 61920 | 60290 |
|---|---|---|---|---|---|
| **age** | 36-40 | 61-65 | 75-80 | 16-20 | 41-45 |
| **sex** | M | F | F | M | F |
| **Calories from saturated fats [kcal] / total calories [kcal]** | 0.1273 | 0.0787 | 0.0992 | 0.1539 | 0.0807 |
| **Low protein-energy-ratio** | 0.0 | 0.0 | 0.0 | 0.0 | 0.0 |
| **Calories from sugar [kcal] / total calories [kcal]** | 0.1603 | 0.1288 | 0.1768 | 0.2163 | 0.3104 |
| **Amount of fiber [g] / total calories [kcal]** | 0.0134 | 0.0106 | 0.0107 | 0.0094 | 0.0182 |
| **Amount of sodium [mg] / total calories [kcal]** | 1.875 | 2.0381 | 1.3206 | 1.6508 | 1.5614 |
| **Low vitamin-C-energy-ratio** | 0.016 | 0.0 | 0.02 | 0.003 | 0.0 |
| **Low iron-energy-ratio** | 0.0 | 0.0 | 0.0 | 0.0 | 0.002 |
| **Calories from beverages [kcal] / weight of beverages [g]** | 0.2899 | 0.0039 | 0.0194 | 0.4108 | 0.1845 |
| **GBS** | 461.0 | 626.0 | 504.0 | 480.0 | 554.0 |
| **Liquids** | BEER, LITE: 1800.0g<br>BEER, LITE: 2160.0g | COFFEE, INSTANT, RECONSTITUTED: 255.0g<br>MILK, FAT FREE (SKIM): 122.0g<br>WATER, TAP: 720.0g<br>TEA, HOT, LEAF, GREEN: 720.0g<br>WATER, TAP: 720.0g<br>WATER, TAP: 720.0g<br>WATER, TAP: 720.0g | WATER, TAP: 207.38g<br>MILK, COW'S, FLUID, SKIM OR NONFAT: 61.25g<br>COFFEE, MADE FROM GROUND, REGULAR: 355.2g<br>ORANGE JUICE, CANNED, BOTTLED OR IN A CARTON: 108.94g<br>WATER, TAP: 1036.88g<br>WATER, TAP: 1110.94g<br>MILK, COW'S, FLUID, SKIM OR NONFAT: 61.25g<br>COFFEE, MADE FROM GROUND, REGULAR: 266.4g | SOFT DRINK, FRUIT-FLAVORED, W/ CAFFEINE: 370.0g<br>MILK, COW'S, FLUID, WHOLE: 1220.0g<br>SOFT DRINK, FRUIT-FLAVORED, W/ CAFFEINE: 370.0g<br>MILK, COW'S, FLUID, WHOLE: 244.0g | CARBONATED WATER, UNSWEETENED (INCL CLUB SODA): 429.2g<br>COFFEE, ESPRESSO: 118.4g<br>COFFEE, MADE FROM GROUND, REGULAR: 1065.6g<br>VODKA: 55.6g<br>CORDIAL OR LIQUEUR: 60.0g<br>LIME JUICE, NS AS TO FORM: 2.56g<br>WINE, TABLE, WHITE: 117.6g<br>CARBONATED WATER, UNSWEETENED (INCL CLUB SODA): 236.8g<br>COFFEE, MADE FROM GROUND, REGULAR: 355.2g |
| **Meat-based dishes** | TACO/TOSTADA W/ BEEF, CHEESE, LETTUCE, TOMATO AND SALSA: 464.0g | DUMPLING, STEAMED, FILLED WITH MEAT, POULTRY, OR SEAFOOD: 18.5g | CHICKEN, RICE & VEG (INCL CAR/DK GRN), TOMATO SAUCE: 715.88g<br>PORK & VEG (NO CAR/DK | CHICKEN OR TURKEY SALAD: 45.5g<br>CHICKEN OR TURKEY SALAD: 91.0g | BEEF STEAK, BROILED OR BAKED, LEAN ONLY: 67.0g<br>SALMON, BAKED OR BROILED: 113.4g |

| | | | | | |
|---|---|---|---|---|---|
| | TUNA SALAD: 85.05g<br>PORK CHOP, BROILED OR BAKED, LEAN ONLY: 94.5g<br>PORK & BEANS: 332.06g | LO MEIN, WITH SHRIMP: 131.25g<br>LO MEIN, WITH PORK: 131.25g<br>LO MEIN, WITH SHRIMP: 131.25g<br>LO MEIN, WITH PORK: 131.25g | GRN, NO POT), TOMATO SAUCE: 217.88g | MEAT LOAF MADE W/ BEEF: 140.0g | |
| **Eggs, Cheese, and Yoghurt** | CHEESE, PROCESSED, AMERICAN/CHEDDAR TYPE: 21.0g | nan | CHEESE, MEXICAN BLEND, REDUCED FAT: 4.71g | YOGURT, FRUIT VARIETY, LOWFAT MILK: 107.19g<br>EGG OMELET OR SCRAMBLED EGG, NO FAT ADDED: 51.0g<br>YOGURT, FRUIT VARIETY, LOWFAT MILK: 141.66g | nan |
| **Grains, Rice, and Noodles** | ROLL, WHITE, SOFT: 36.0g<br>CRACKER, SNACK: 27.0g | nan | OATMEAL, COOKED, REGULAR, NO FAT ADDED: 307.13g<br>TORTILLA, CORN: 48.07g<br>TORTILLA, FLOUR (WHEAT): 33.18g<br>OATMEAL, COOKED, INSTANT, NO FAT ADDED IN COOKING: 307.13g<br>TORTILLA, FLOUR (WHEAT): 45.0g<br>TORTILLA, FLOUR (WHEAT): 135.0g<br>CRACKER, ANIMAL (INCL ARROWROOT COOKIE): 32.0g<br>ROLL, SWEET, NO TOPPING, MEXICAN (PAN DULCE): 76.0g | BREAD, WHEAT OR CRACKED WHEAT: 28.0g<br>BREAD, WHOLE WHEAT, NS AS TO 100%: 52.0g | CRACKERS, SALTINE, WHOLE WHEAT: 30.0g |
| **Snacks** | REESE'S PEANUT BUTTER CUPS: 34.0g<br>CHEWING GUM, NFS: 5.6g<br>POPCORN, POPPED IN OIL, UNBUTTERED: 22.0g<br>RAISINS (INCLUDE CINNAMON-COATED RAISINS): 72.5g | nan | ICE CREAM, REGULAR, NOT CHOCOLATE: 83.25g<br>ICE CREAM, REGULAR, NOT CHOCOLATE: 180.0g<br>CHEESECAKE, DIET: 28.93g | WHITE POTATO, CHIPS, RESTRUCTURED: 10.0g | CHOCOLATE CANDY, SWEET OR DARK: 15.2g<br>SORBET, FRUIT, NONCITRUS FLAVOR: 112.5g<br>SORBET, FRUIT, NONCITRUS FLAVOR: 50.0g<br>CHOCOLATE CANDY, SWEET OR DARK: 14.18g |
| **Fruits** | nan | GRAPEFRUIT, RAW: 64.0g<br>PEAR, RAW: 28.0g<br>APPLE, RAW: 45.5g | BANANA, RAW: 101.0g | FRUIT JUICE BAR, FROZEN, FLAVOR OTHER THAN ORANGE: 74.0g | BLACKBERRIES, RAW: 126.0g<br>STRAWBERRIES, RAW: |

| | | | | | |
|---|---|---|---|---|---|
| | | | | | 189.0g<br>APPLE, RAW: 223.0g<br>STRAWBERRIES, RAW: 144.0g |
| **Vegetables, Beans, and Lentils** | ONIONS, MATURE, RAW (INCLUDE RED ONIONS, NFS): 40.0g<br>LETTUCE, RAW (INCLUDE LETTUCE, NFS): 32.0g<br>TOMATOES, RAW: 91.0g<br>CORN, YELLOW, COOKED, FROM CANNED, FAT ADDED: 84.5g<br>TOMATOES, RAW: 27.0g<br>LETTUCE, RAW (INCLUDE LETTUCE, NFS): 25.0g | CARROTS, RAW, SALAD WITH APPLES: 224.44g | SWEETPOTATO, CANDIED: 98.0g<br>PINTO, CALICO/RED/MEX BEAN, DRY, COOKED, NO FAT: 86.5g | nan | WHITE POTATO, ROASTED, FAT ADDED: 166.69g<br>CARROTS, COOKED, FROM FRESH, FAT ADDED: 52.83g<br>SQUASH, SUMMER, COOKED, FROM FRESH, FAT ADDED: 121.41g<br>BEANS, STRING, GREEN, COOKED, FROM FRESH, FAT ADDED: 42.66g<br>SPINACH, RAW: 45.0g<br>LETTUCE, RAW: 41.25g<br>TOMATOES, RAW: 135.0g<br>PEPPERS, PICKLED: 16.88g<br>HUMMUS: 61.5g |
| **Nuts** | nan | nan | PEANUT BUTTER: 16.0g<br>PEANUT BUTTER: 10.67g<br>PEANUTS, NFS: 8.31g | nan | nan |
| **Other** | SALSA, RED, CKD, NOT HOM (INCL TACO, CREOLE, PICANTE SAUCES): 35.6g | nan | nan | nan | SUGAR, WHITE, GRANULATED OR LUMP: 2.8g<br>ITALIAN DRESSING, W/ VINEGAR & OIL: 58.8g |
